# Concordance between Infectious Disease Society of America Criteria for Urinary Tract Infection Testing and Treating vs. Actual Emergency Department Practice

**DOI:** 10.1101/2024.10.01.24314581

**Authors:** Tarek Harhash, Yi-Ru Chen, Sahit Dokku, Demetra Menoudakos, Keyleen Argueta, Musa Almiggaber, Chelsea Rampersad, Mark Richman

## Abstract

**Introduction:** Symptomatic and asymptomatic urinary tract infections (UTIs) are common. The Infectious Disease Society of America (IDSA) discourages testing and treating ASB other than in pregnant women during routine obstetric visit screening and patients undergoing urologic procedures with expected mucosal bleeding. Unnecessary urinalysis (UA) and inappropriate antibiotic use persist in Emergency Departments (EDs). This study aims to evaluate UA testing and antibiotic treatment patterns for ASB in an urban ED, assessing adherence to IDSA guidelines and setting baseline rates for an educational intervention to align testing and treatment with IDSA guidelines.

**Methods:** We conducted a 15-month study to assess adherence to IDSA guidelines for proper UTI screenings and prescribing. We reviewed records of 50 adult patients who had a UA at the Long Island Jewish Medical Center ED to determine whether they met IDSA criteria for UA testing and appropriate antibiotic use. Patients with sepsis or other conditions requiring empiric antibiotics were excluded. We performed a univariate analysis to describe the population and factors associated with treating urinalysis findings. Statistical significance was set at p <0.05.

**Results:** Sixty-four percent of patients were asymptomatic and 36% were symptomatic. None of the asymptomatic patients met IDSA criteria for UA testing. Symptomatic patients were nearly-statistically more likely than asymptomatic patients to have a positive UA (72.2% vs. 43.8%, p = 0.06), and were more-often prescribed antibiotics for a positive UA (61.5% vs. 14.3%; p = 0.0128). They were also more-often prescribed antibiotics for a negative UA (20.0% vs. 0%; p = 0.05).

**Discussion:** The study findings revealed significant discordance between IDSA guidelines and current ED practices, with 64% of UA tests deemed unnecessary for ASB patients. These results align with previous studies highlighting the prevalence of over-testing with UAs and over-treatment of ASB. Unnecessary testing and inappropriate antibiotic treatment lead to increased costs and risks of antibiotic resistance, adverse drug events, and Clostridium difficile infection. This study highlights the necessity for planned educational initiatives to reduce unnecessary UAs and treatment and improve adherence to evidence-based guidelines.

## Introduction

Urinary tract infections (UTIs) are one of the most-commonly diagnosed infections worldwide, with over 400 million cases and over 200,000 deaths in 2019.^1^ UTI incidence and complications vary dramatically with gender and age. Females have an estimated 60% lifetime incidence of contracting a UTI, and men a 10% lifetime incidence.^2^ Increased risk of infections among women is due to anatomical differences, as females have a shorter urethra closer to the anus, making it easier for gastrointestinal bacteria to ascend to the bladder. Additional risk factors for UTIs are neurologic and immune deficiency conditions, such as diabetes and chemotherapy.^3^ Lower tract UTIs involve the bladder only, while upper tract infections involve the ureters and kidneys. Common UTI symptoms include cloudy urine, dysuria, flank pain, foul-smelling urine, passing small amounts of urine, supra-pubic pain, urinary frequency, and urinary urgency.^4^

UTIs are diagnosed by a urinalysis (UA) showing bacteria under microscopy, or a urine culture (UCx) growing >100,000 colony-forming units (CFUs). A UTI can be suggested by inflammatory changes in the urine (eg, white blood cells (WBCs) or leukocyte esterase (LE), or by bacterial metabolic byproducts (eg, nitrite). This has utility in venues such as the Emergency Department (ED) if bacteria are not seen by microscopy and culture has not yet resulted.

UTIs can be symptomatic or asymptomatic. The prevalence of asymptomatic bacteriuria (ASB) in patients 65 to 80 years old is greater than 15%, and this rises to 40-50% in those over 80 years old.^5^ ASB Symptomatic patients should be treated.^6^ UTIs can be further categorized as “treatable” or “untreatable,” according to the Infectious Disease Society of America (IDSA). There is no evidence to support treating ASB except in pregnant patients and patients before invasive urologic procedures (eg, trans-urethral resection of the prostate (TURP)) in which mucosal bleeding is expected. While screening for ASB is part of routine obstetrics care,^7^ there is no role for screening for ASB in a pregnant patient in the ED. The following groups among whom UAs are commonly sent despite lacking UTI symptoms are advised against undergoing screening or treatment for ASB: premenopausal, non-pregnant women; diabetic patients; functionally impaired older adults, assisted living; elderly, institutionalized subjects; kidney transplant recipients; solid organ transplant recipients; elective neurologic surgery; elective orthopedic surgery; persons with spinal cord injury; or catheterized patients while the catheter remains in situ.^6^

However, there are circumstances in which patients with ASB are not included or excluded from this list. This “gray area” includes functionally-impaired elderly adults in long-term care facilities, as well as cognitively-impaired patients with non-localizing symptoms (eg, altered mental status, lethargy, unsteady gait/falls).^6^ In the absence of typical UTI symptoms, the IDSA recommends searching for another reason for non-localizing symptoms, instead of sending a UA or UCx. Identifying ASB in such patients should not reflexively lead to treatment, as alternate etiologies may better explain their signs and symptoms. However, in EDs, where providers manage multiple patients at one time and, in most cases, a confirmed or highly-likely diagnosis is expected prior to discharge, it is reasonable to expect many providers to send a UA and/or UCx on “gray area” patients.

As suggested above, checking UA and UCx should not change management (ie, shouldn’t lead to antibiotic prescribing) in many populations with ASB (eg, diabetic patients, elderly, institutionalized). Nonetheless, inappropriate testing and treating populations for ASB is common. One study of veterans in long-term care facilities examining almost 20,000 patients over 5 years showed a 64.6% incidence of potentially-suboptimal antibiotic prescriptions.^8^ Another study, investigating two insurance plans over 10 years, estimated $48 million was spent on inappropriate urinalyses and over $4 million in additional spending on inappropriate antibiotic prescriptions.^9^ Multiple randomized control trials have demonstrated that treatment of ASB with antibiotics offer no clinical benefits for adults, as well as potentially causing significant harms.^10,11,6,12^

Harms of treating ASB include increased risk for future symptomatic UTI, complicated UTIs, pyelonephritis, sepsis,^13^ and antibiotic resistance.^6^ This latter problem is of particular public health importance, given the decline in the creation of new antibiotics.^14^ Additional complications include increasing ED visits or hospitalization, adverse drug events, and Clostridium difficile infection.

The psychology of practitioners makes it difficult not to treat a positive result, even when evidence argues against such treatment.^15^ For that reason, antibiotic stewards advocate not testing patients in whom there is no indication for treatment.^16,17^ Given lack of benefit, and several potential harms, associated with unnecessary antibiotic treatment in ASB patients, it is important both to individual patients and the public’s health that practitioners adopt more evidence-based urine testing practices. Such behavior change often starts with demonstrating to practitioners their current practice patterns^18^ as many have the “Lake Wobegon” effect that it is others, but not they, who are practicing contrary to the evidence.^19^

Our experience as academic Emergency Medicine (EM) physicians at an urban tertiary care hospital has been that many ASB patients undergo unwarranted urine testing and treatment for infection. This study aims to evaluate our ED’s current ordering and treatment patterns for patients with ASB. If the data confirm our experience, we will use the results to support an initiative toward more IDSA-guideline-concordant ASB testing and treatment behavior.

## Methods

Northwell Health is a healthcare system with 23 hospitals and 890 outpatient facilities across New York. Long Island Jewish Medical Center (LIJMC) is a 583-bed tertiary-care academic hospital, treating a diverse range of racial and socioeconomic populations. The adult ED at LIJMC has approximately 100,000 patients per year. From June 28, 2023, through September 2, 2023, we retrospectively collected data from a random sample of 50 LIJMC ED patients ≥18 years old who had a UA. The researchers reviewed charts to determine whether the patient was symptomatic or not, whether they met IDSA criteria for ordering a UA (plan for emergent invasive urologic procedure with expected mucosal bleeding, such as a stent for renal stone), and whether they were treated with antibiotics. Patients were excluded if they manifested systemic inflammatory response syndrome (SIRS) or sepsis according to the Surviving Sepsis Campaign^20^ for which empiric antibiotic perceptions are appropriate and with which all our ED providers are familiar, as sepsis guideline adherence is a departmental strategic initiative.

The operational definition of symptoms for which a patient might have a symptomatic UTI (and, therefore, for which a UA, urine culture, and empiric antibiotics were warranted) were: cloudy urine, dysuria, fever, flank pain, urine frequency, penile discharge, suprapubic pain, vaginal itching, vaginal discharge, and unusual odor. A patient was determined to have a UTI (whether or not it was a treatable UTI) if any of the following criteria were present on urinalysis: bacteria; nitrite; or >10 WBCs plus moderate-to-large leukocyte esterase in the absence of bowel pathology such as appendicitis or diverticulitis (as such nearby non-urinary tract pathology might cause reactive pyuria (sterile pyuria)). As this study focused on ED management of UTIs, we did not include data regarding urine cultures, which take 24-48 hours to result (ie, long after a patient is discharged from the ED).

We performed a univariate analysis to describe the population and factors associated with treating urinalysis findings. Statistical significance was set a priori at p <0.05. This study was deemed by the Northwell Health Institutional Review Board (HSRD19-0392) not to be research, but, instead, to be a quality improvement project to assess whether education works in helping staff adhere to evidence-based guidelines.

## Results

Among the 50 patients (all of whom had a UA), 22% (11) were male and 78% (39) were female. (**Table 1**). Males comprised 33.3% of symptomatic patients vs. 15.6% of asymptomatic patients. The median patient age of all patients was 60 years (symptomatic: median = 54 years; asymptomatic: median = 71.5 years).

| **Clinical variable** | **Category** | **Count** | **%** |
| --- | --- | --- | --- |
| Age (years) | 18-30 | 7 | 14% |
|  | 31-50 | 11 | 22% |
|  | 51-70 | 10 | 20% |
|  | >70 | 22 | 44% |
| Gender | Male | 11 | 22% |
|  | Female | 39 | 78% |

Vital signs that might have prompted a UA (eg, temperature ≥38.4 Celcius, systolic blood pressure <100 mmHg, heart rate >100 beats/minute) were similar between symptomatic and asymptomatic patients (**Table 2**). Of all patients, 64% (32) were asymptomatic and 36% (18) were symptomatic. None of the asymptomatic patients met IDSA criteria for testing (eg, none were to receive a near-term invasive urologic procedure, and there is no role for screening for asymptomatic bacteriuria in pregnant ED patients). Nearly three-quarters (72.2%) of symptomatic patients had a positive UA, and 61.5% of these received antibiotics. In contrast, 43.8% of asymptomatic patients had a positive UA, and 14.3% of these received antibiotics. (**Table 3, Figure 1**) Five symptomatic patients were not prescribed an antibiotic.

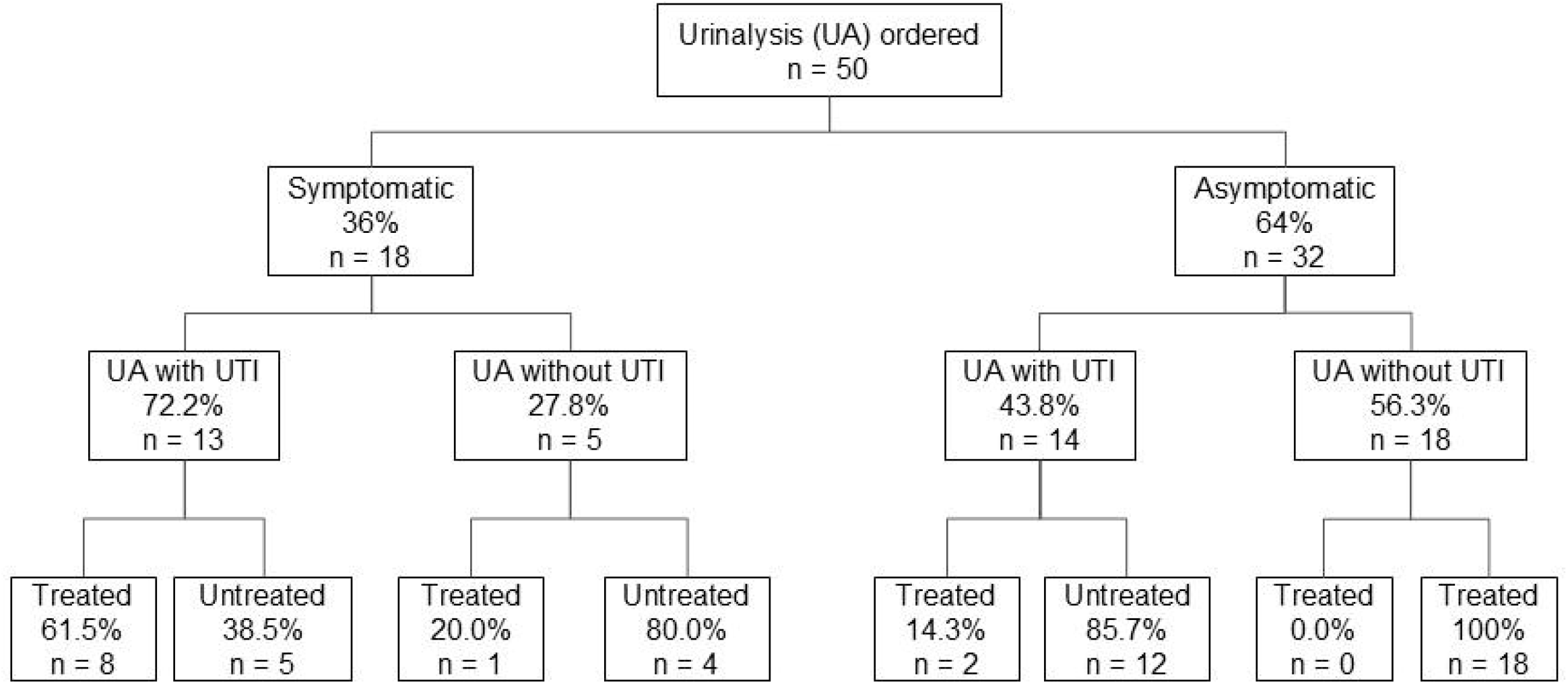

| **Vital Sign** | **Symptomatic** | **Asymptomatic** | **P-Value** |
| --- | --- | --- | --- |
| Temperature $\geq 38.4$ | 5.60% | 0.00% | 0.1806 |
| SBP $< 100$ | 5.60% | 18.80% | 0.2018 |
| HR $> 100$ | 5.60% | 3.10% | 0.6682 |

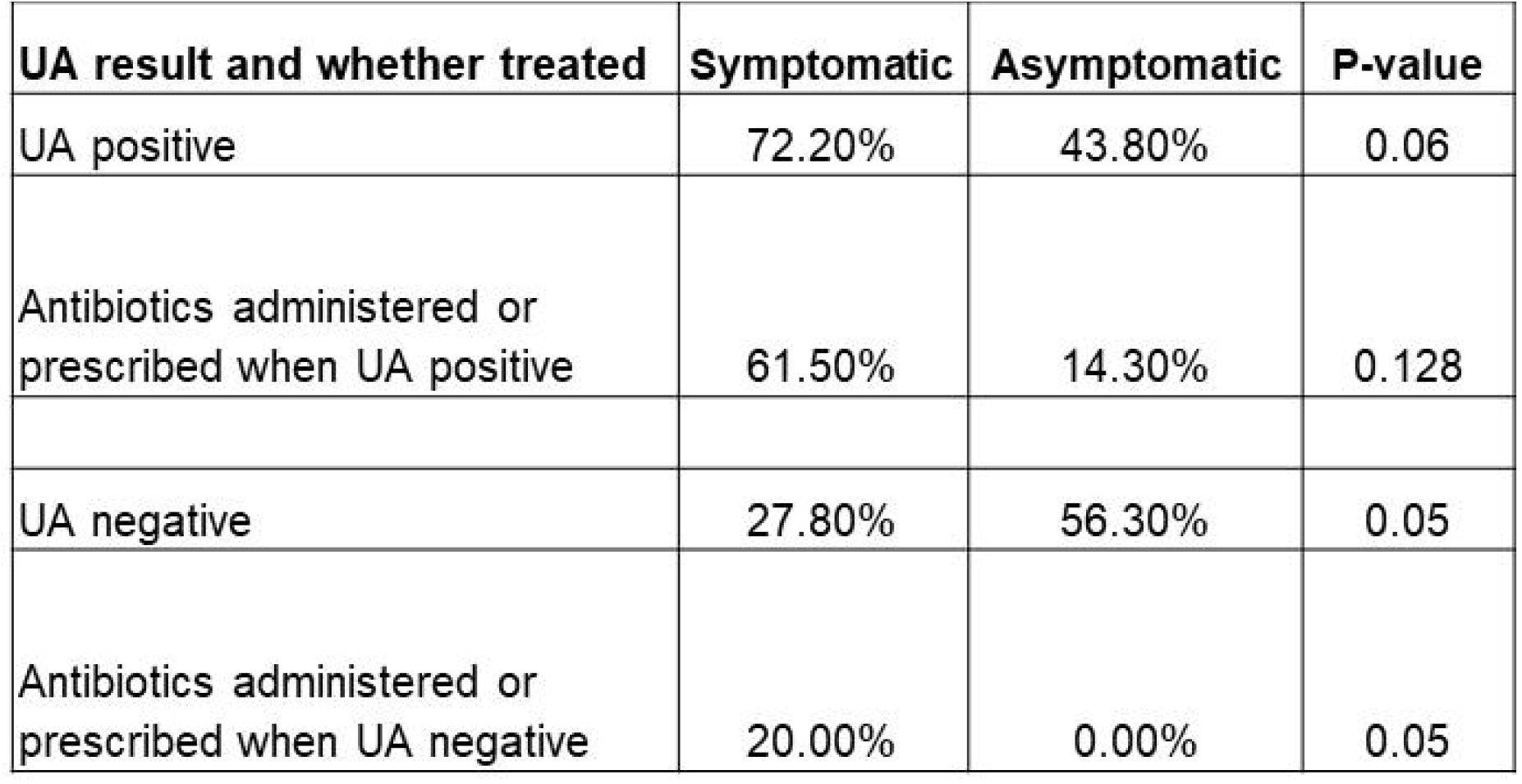

In 54% (27 of 50) of cases, the UA met criteria for a UTI (eg, nitrite positive). Symptomatic patients were nearly-statistically more likely than asymptomatic patients (72.2% vs. 43.8%, p = 0.06) to have a positive UA, and were more-often prescribed antibiotics for a positive UA (61.5% vs. 14.3%; p = 0.0128). They were also more-often prescribed antibiotics for a negative UA (20.0% vs. 0%; p = 0.05).

## Discussion

This 50-patient ED study found that, in nearly ⅔ of patients on whom a UA was sent, the UA was unwarranted per IDSA guidelines or vital signs that might justify sending a UA. These findings show significant discordance between ED practice and IDSA guidelines. The IDSA Guidelines in Nicolle et al. 2019 emphasize that testing and treatment of ASB should be highly-selective, recommended only for pregnant women and patients undergoing invasive urologic procedures.^6^ A study published by *IM Matters* highlights how clinical misjudgments frequently contribute to the overprescription of antibiotics for ASB.^21^ Over-treatment has been well-documented in the literature, with significant concerns about causing antimicrobial resistance, adverse drug events (eg, gastrointestinal discomfort), Clostridium difficile infection, and disruption of the normal microbiota.^13^ In our study, 14.3% of asymptomatic patients with a positive UA received antibiotics when, per IDSA guidelines, none should have. This is consistent with prior studies that found over-testing and over-treating to be common.^22,23^ Such studies include a 2019 study that analyzed patients’ outcomes across Michigan over two years. The Michigan Hospital Medicine Safety Consortium conducted a retrospective cohort study, including 2,733 patients from 46 hospitals, to assess risk factors associated with antibiotic treatment of ASB and evaluate patient outcomes. The study discovered that 83% of patients with ASB received antibiotics.^13^ Another study investigated unnecessary antibiotic treatment in a large population of hospitalized patients with ASB; nearly 70% of patients with ASB without SIRS criteria were treated with antibiotics.^24^ The safety of withholding antibiotics in ASB was validated in another study that found no significant differences in mortality, readmissions, or ED visits between ASB patients treated with antibiotics and those who were not.^13^ In this current study, 5 symptomatic patients with a positive UA at the index visit captured in our data did not receive antibiotics. Chart review revealed that four of the five (80%) were already taking antibiotics for a UTI; the symptoms captured in our study were deemed due to an inadequate number of days taking the antibiotic from their prior visit.

However, there are situations in which treating patients despite a negative UA result is acceptable, as a diagnosis can be made clinically. A literature review by the American Academy of Family Physicians (AAFP) indicated patients can be treated based on their clinical factors and symptoms, even with a negative UA (ie, UTI can be a clinical diagnosis). Urinary frequency, urgency, dysuria, and suprapubic pain were all predictive of UTI, as indicated by a positive urine culture, despite a negative UA.^24^ Furthermore, a review of 464 studies examining the role of history-taking in diagnosing UTIs in women found that those with dysuria, increased urinary frequency, no risk factors for complicated infection, and no vaginal discharge had a 90% probability of having a UTI.^24^ The study advocated treating UTI based on symptoms alone. Another prospective study of 490 men with symptoms of a UTI indicated that symptoms of dysuria and urgency were significantly associated with positive urine cultures.^25^

### Limitations

This study was based on data from a single ED, which might limit generalizability. Furthermore, all data collected pertaining to this study was only in the timeframe between June 2023-September 2023; testing and treatment patterns may have differed during other timeframes. In addition, the study sample size was small; however, this was intended to obtain baseline data in preparation for an educational intervention. Finally, we performed only a univariate analysis that did not account simultaneously for confounding patient, provider, or operational variables. Such variables include patient preference for antibiotic treatment, provider awareness of IDSA guidelines, and ED overcrowding (which might impact whether the provider evaluates a patient before ordering a urinalysis, as providers often order studies before seeing a patient).^26^

## Conclusion

Unnecessary urinalyses continue to be commonly-ordered, and asymptomatic bacteriuria often treated, in contradiction to Infectious Disease Society of America guidelines. Such unwarranted practices not only pose a financial burden, but also significantly increase the risks of antibiotic side effects and resistance. Future clinical education should focus on reducing unnecessary urinalyses and urine cultures in accordance with IDSA guidelines. This study provides a baseline for a planned educational intervention to emphasize the IDSA guidelines. Post-education, we will re-evaluate urinalysis testing and treatment rates.

## Data Availability

All data produced in the present study are available upon reasonable request to the authors

